# High-Risk Endometrial Cancer Assessed by Immediate Intraoperative Frozen Sections of Sentinel Lymph Nodes – A Retrospective Study

**DOI:** 10.1101/2021.06.22.21258922

**Authors:** Sarah E Miller, Mahkam Tavallaee, Malte Renz, Ann K Folkins, Amer Karam

## Abstract

While sentinel lymph node (SLN) sampling has been established for low-risk endometrial cancer, few data exists on high-risk histologies. This study aims to measure the accuracy of immediate intraoperative SLN biopsy with frozen section in high-risk endometrial cancer. Patients diagnosed with endometrial cancer of high-grade histology (grade 3 endometrioid, clear cell, serous, carcinosarcoma, de- or undifferentiated histology) between 2014 and 2019 at a single institution who underwent SLN mapping, followed by pelvic lymphadenectomy with or without para-aortic lymphadenectomy were included. SLNs were assessed intraoperatively using multiple frozen sections and H&E staining. Lymph node metastases detected by SLN biopsy were compared with complete lymphadenectomy specimens.

35 patients with high-grade endometrial cancer histology underwent SLN mapping followed by lymphadenectomy. In 34 of 35 (97%) of these patients mapping with at least one SLN was successful. Positive SLNs were identified in 7/34 patients (20.6%). There were no patients who had positive lymph nodes on complete lymphadenectomy without a positive SLN, resulting in 100% sensitivity, and 0% false-negative rate.

SLN mapping using intraoperative frozen sections in high-risk endometrial cancer demonstrated 100% sensitivity and 0% false-negative rate, provides immediate feedback on successful SLN mapping and valuable intraoperative information on the disease status guiding the intraoperative decision for completion lymphadenectomy.

**Highlights:**

- Intraoperative SLN assessment in high-grade endometrial cancer by frozen section.
- 100% sensitivity and 0% false-negative rate compared to complete lymphadenectomy.
- Protocol provides immediate intraoperative feedback on successful SLN mapping.
- Protocol helps inform surgeon’s decision to proceed with complete lymphadenectomy.

## Introduction

In 1987, the GOG 33 trial results defined risk factors of extra-uterine spread, specifically lymph node metastases, for clinically uterine confined endometrial cancers [1]. Subsequently, in 1988, the FIGO staging for endometrial cancer was revised from a clinical staging to a surgical staging system that includes surgical lymph node assessment [2]. Randomized control trials, however, did not show any progression-free or overall survival benefit from complete lymphadenectomy in clinically early-stage uterine cancer [3, 4]. Sentinel lymph node injection and biopsy was introduced in order to reduce surgical morbidity associated with lymphadenectomy, particularly leg lymphedema, while maintaining the prognostic information gained from surgical staging and help guide adjuvant treatment decisions. While sentinel lymph node injection and biopsy has become an accepted approach for low-risk endometrial cancer, little data exists on endometrial cancer of high-risk histologies [5]. As per NCCN guidelines, sentinel lymph node injection and mapping may be used for high-risk histologies as well. These guidelines also state that sentinel lymph nodes should be processed using ultrastaging which includes serial sectioning and cytokeratin immunohistochemistry but standard protocols are not available [6]. In addition, the clinical significance of isolated tumor cells (ITC) defined as tumor deposits ≤ 0.2 mm and ≤ 200 tumor cells remains uncertain.

Previously published retrospective analyses of high-risk endometrial cancer patients have reported on sentinel lymph node injection and mapping combined with ultrastaging and immunohistochemistry. The reported false-negative rates range from 4.2% to 22% [7-11]. We report the application of our previously published protocol for immediate intraoperative sentinel lymph node assessment by frozen section [12] for high-risk endometrial cancers. In high-risk endometrial cancer the immediate intraoperative feedback on successful SLN mapping and the disease status is especially important and may guide the intraoperative decision for a completion lymphadenectomy.

## Material and Methods

### Patients

Electronic medical records from 2014 to 2019 were reviewed, and consecutive patients with high-grade endometrial cancer (grade 3 endometrioid, clear cell, serous, carcinosarcoma, de- or undifferentiated histology) were evaluated for inclusion. Inclusion criteria were: high-grade histology on final pathology of the hysterectomy specimen, age older than 18 years, patients who underwent SLN injection and mapping followed by complete lymphadenectomy. The study was approved by our Institutional Review Board (IRB).

### Procedure

Following successful induction of anesthesia, the cervix was injected in four quadrants with 0.5 mL of a 0.5 mg/ mL indocyanine green solution. The Intuitive da Vinci Si or Xi surgical robotic systems were used for all robotic-assisted laparoscopic surgeries. Efferent lymphatic vessels and sentinel lymph nodes were visualized using near-infrared imaging, and sentinel lymph nodes were sent to pathology for intraoperative evaluation via frozen section. Subsequent complete pelvic lymphadenectomy was then performed according to the Gynecologic Oncology Group surgical handbook [13]. In agreement with current NCCN guidelines, para-aortic lymph node dissection was performed when risk factors were identified and at the discretion of the physician. The study was IRB approved and patients were consented for sentinel lymph node injection and biopsy and lymph node dissection.

### Pathology

Following dissection and removal, sentinel lymph nodes sent for intraoperative analysis were either partially or entirely submitted for microscopic examination at the discretion of the pathologist. Lymph nodes that were partially submitted were assessed intraoperatively by evaluating 2 hematoxylin and eosin (H&E) stained sections, with additional permanent sections (2-4) evaluated post-operatively. In contrast, entirely submitted lymph nodes were solely assessed intraoperatively by evaluating 4 or more H&E stained sections at levels evenly distributed levels in the tissue block. A lymph node metastasis was defined as the presence of tumors cells of any number or size ranging from isolated tumor cells to macro-metastases.

### Statistics

Successful sentinel lymph node mapping was calculated as the number of patients with at least one detected SLN divided by the total number of patients who underwent mapping. We also calculated the rate of bilateral sentinel lymph node mapping. False negative cases were defined as patients whose sentinel lymph node sampling was negative for disease but who had disease detected on subsequent pelvic lymphadenectomy. Risk factors for sentinel and non-sentinel lymph node metastases were evaluated using Fisher’s Exact test. A p-value of < 0.05 was considered statistically significant. Data regarding intraoperative and postoperative complications, such as urinary and nerve injuries as well as lymphoceles and lymphedema, was derived from operative reports and postoperative follow up notes.

## Results

Between April 2014 and September 2019, 35 patients with high-risk endometrial cancer underwent sentinel lymph node injection and sampling followed by complete lymphadenectomy by one surgeon at Stanford University. The mean age was 66 +/- 9 years and the mean body mass index 29 +/- 2 kg/m^2^.

Final pathological diagnosis and FIGO stage of the 36 patients are outlined in Table 1. 27 out of 35 patients (77%) had endometrial cancers confined to the uterus on final pathology (Stage IA or IB).

**Table 1:**
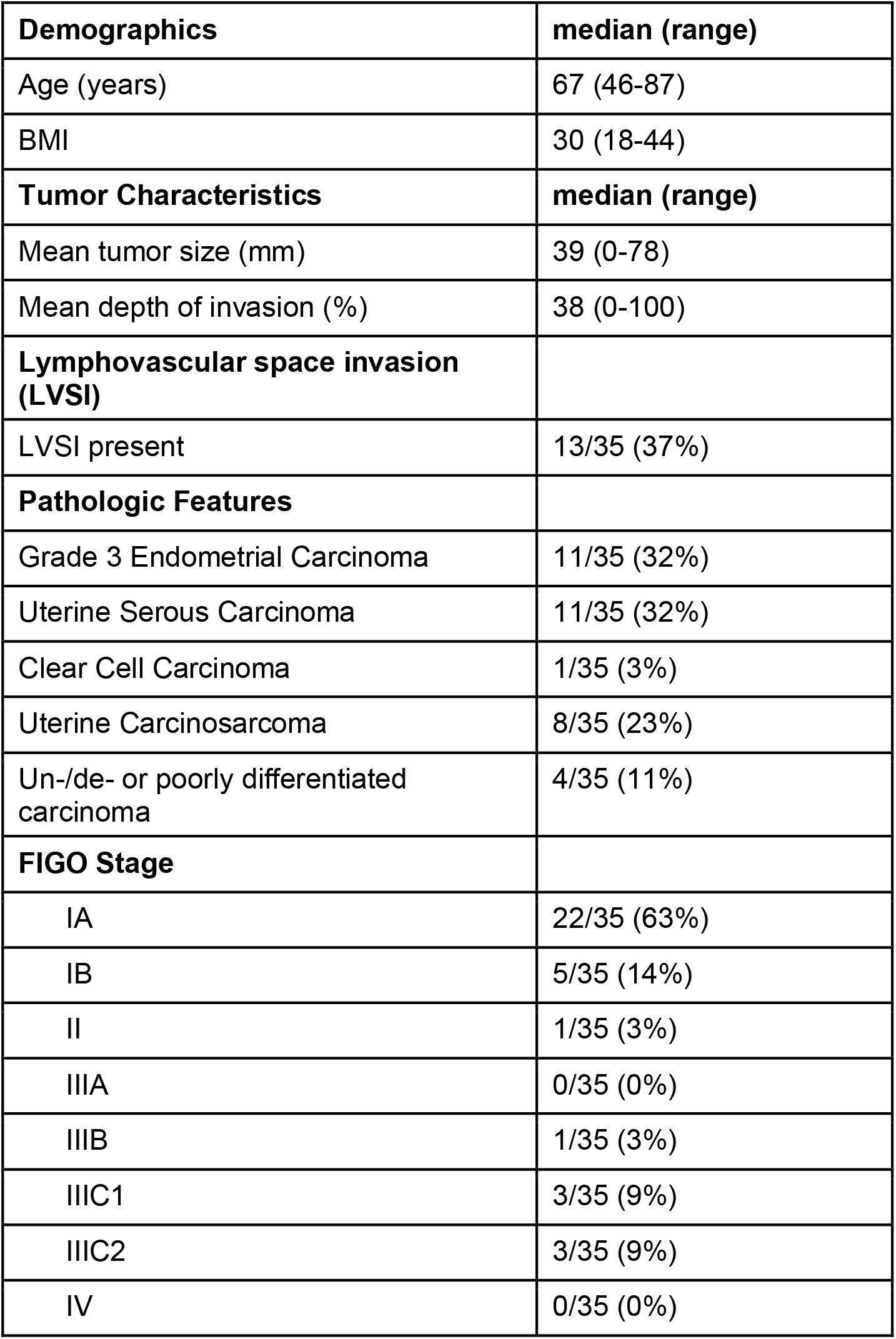
Patient demographics, tumor characteristics, pathologic features and postoperative stage.

| **Demographics** | **median (range)** |
| --- | --- |
| Age (years) | 67 (46-87) |
| BMI | 30 (18-44) |
| **Tumor Characteristics** | **median (range)** |
| Mean tumor size (mm) | 39 (0-78) |
| Mean depth of invasion (%) | 38 (0-100) |
| **Lymphovascular space invasion (LVSI)** |  |
| LVSI present | 13/35 (37%) |
| **Pathologic Features** |  |
| Grade 3 Endometrial Carcinoma | 11/35 (32%) |
| Uterine Serous Carcinoma | 11/35 (32%) |
| Clear Cell Carcinoma | 1/35 (3%) |
| Uterine Carcinosarcoma | 8/35 (23%) |
| Un-/de- or poorly differentiated carcinoma | 4/35 (11%) |
| **FIGO Stage** |  |
| IA | 22/35 (63%) |
| IB | 5/35 (14%) |
| II | 1/35 (3%) |
| IIIA | 0/35 (0%) |
| IIIB | 1/35 (3%) |
| IIIC1 | 3/35 (9%) |
| IIIC2 | 3/35 (9%) |
| IV | 0/35 (0%) |

In 34 of the 35 patients (97%), at least one lymph node was successfully mapped (Table 2). In 26 of the 35 patients (74%), bilateral mapping was successful. While complete pelvic lymphadenectomy was done in all patients, para-aortic lymphadenectomy was concurrently performed in 21 out of 35 patients (60%). The median number of sentinel nodes removed was 2 (0-13) and the mean number of total nodes removed during complete lymphadenectomy was 24 +/- 10.

**Table 2:**
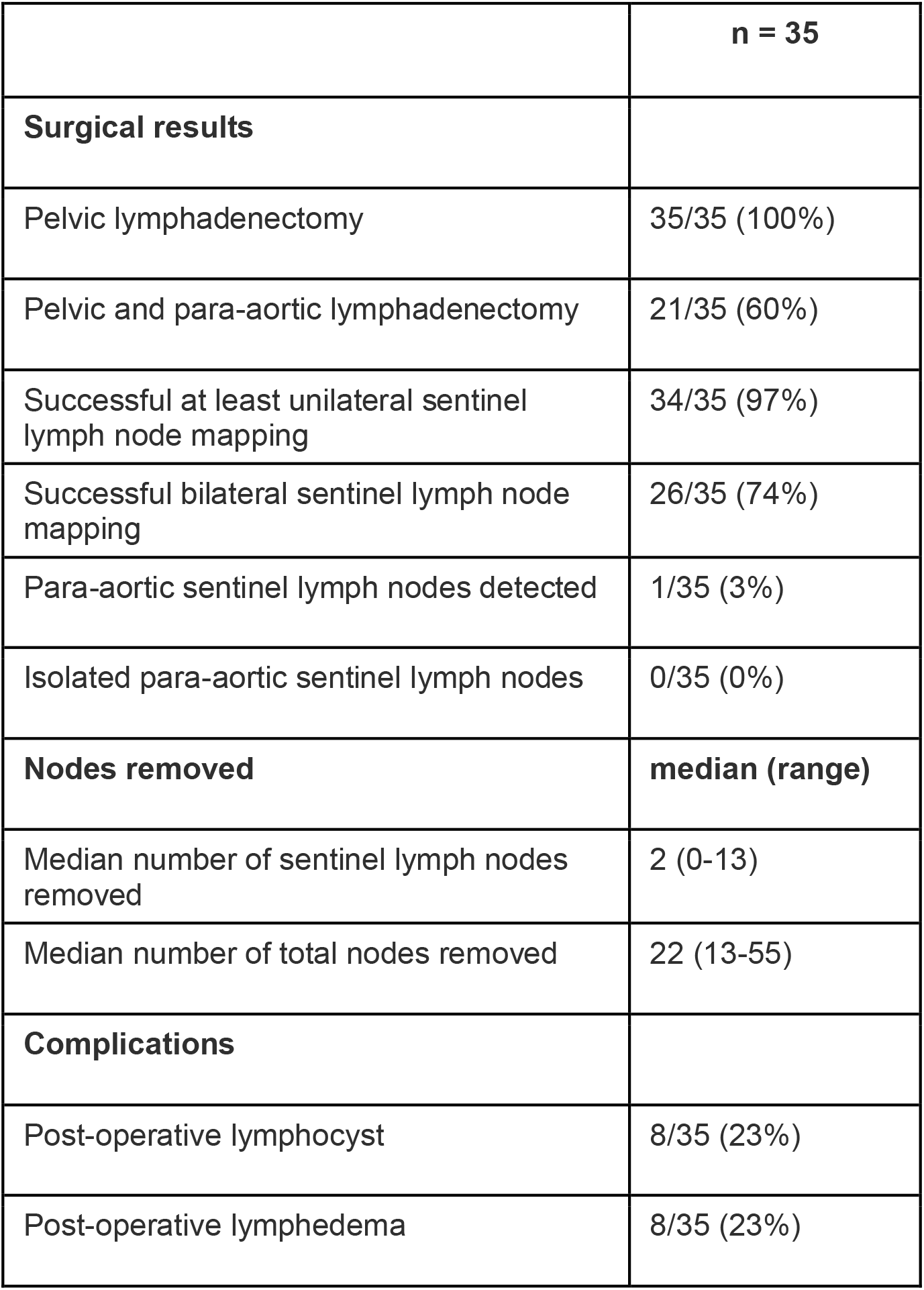
Surgical results and complications in patients with lymphadenectomy.

|  |  |
| --- | --- |
|  | **n = 35** |
| **Surgical results** |  |
| Pelvic lymphadenectomy | 35/35 (100%) |
| Pelvic and para-aortic lymphadenectomy | 21/35 (60%) |
| Successful at least unilateral sentinel lymph node mapping | 34/35 (97%) |
| Successful bilateral sentinel lymph node mapping | 26/35 (74%) |
| Para-aortic sentinel lymph nodes detected | 1/35 (3%) |
| Isolated para-aortic sentinel lymph nodes | 0/35 (0%) |
| **Nodes removed** | **median (range)** |
| Median number of sentinel lymph nodes removed | 2 (0-13) |
| Median number of total nodes removed | 22 (13-55) |
| **Complications** |  |
| Post-operative lymphocyst | 8/35 (23%) |
| Post-operative lymphedema | 8/35 (23%) |

Table 3 shows the total number of sentinel lymph nodes successfully mapped and the number of positive sentinel lymph nodes in this series. 118 nodes were successfully identified and removed; 63 from the right side and 55 from the left. Predominant locations of the detected sentinel lymph nodes were in the external iliac area with 58.5% (69/118) and the obturator space with 36.4% (43/118). In this series, no isolated para-aortic sentinel lymph nodes were detected.

**Table 3:** Numbers of positive SLN/successfully mapped SLN.

| SLN site |  |
| --- | --- |
| Right Para-aortic | 0/1 |
| Right common iliac | 0/4 |
| Right external iliac | 3/35 |
| Left external iliac | 2/34 |
| Right obturator | 4/22 |
| Left obturator | 1/21 |
| Right parametrial | 0/1 |

Sensitivity and specificity data were calculated from the 34 patients in which at least one sentinel lymph node was successfully mapped (Table 4). Both, sensitivity and specificity were 100%. No false-negative sentinel lymph nodes were detected. 7 out 34 patients (20.6%) showed positive sentinel lymph nodes. 3 out of the 7 patients (42.8%) with positive sentinel lymph nodes also had positive non-sentinel lymph nodes. All 3 patients with positive sentinel and non-sentinel lymph nodes had positive para-aortic non-sentinel lymph nodes as well. In one patient, the para-aortic non-sentinel lymph nodes were positive even in the absence of positive pelvic non-sentinel lymph nodes. Clinical and histological characteristics and anatomical locations of the positive sentinel lymph nodes are described in Supplementary Table 1 and 2. Table 5 displays risk factors associated with lymph node metastases. In this series, a statistically significant correlation was noted between lymph node metastases and lymphovascular space invasion (p=0.0059), depth of myometrial invasion (p=0.0238), and age (p=0.0238).

**Table 4:** Sensitivity and specificity data.

|  | **True Positive**<br>**Nodes** | **True Negative**<br>**Nodes** |  |
| --- | --- | --- | --- |
| **Positive**<br>**SLN** | 7 | 0 | PPV 7/7<br>(100%) |
| **Negative**<br>**SLN** | 0 | 27 | NPV<br>27/27<br>(100%) |
|  | Sn 7/7 (100%) | Sp 27/27 (100%) | ACC<br>34/34<br>(100%) |
|  | FNR 0/7 (0%) | FPR 0/27 (0%) |  |

Sn = Sensitivity, Sp = Specificity, FNR = False Negative Rate, FPR = False Positive Rate, PPV = Positive Predictive Value, NPV = Negative Predictive Value, ACC = Accuracy

**Table 5:**
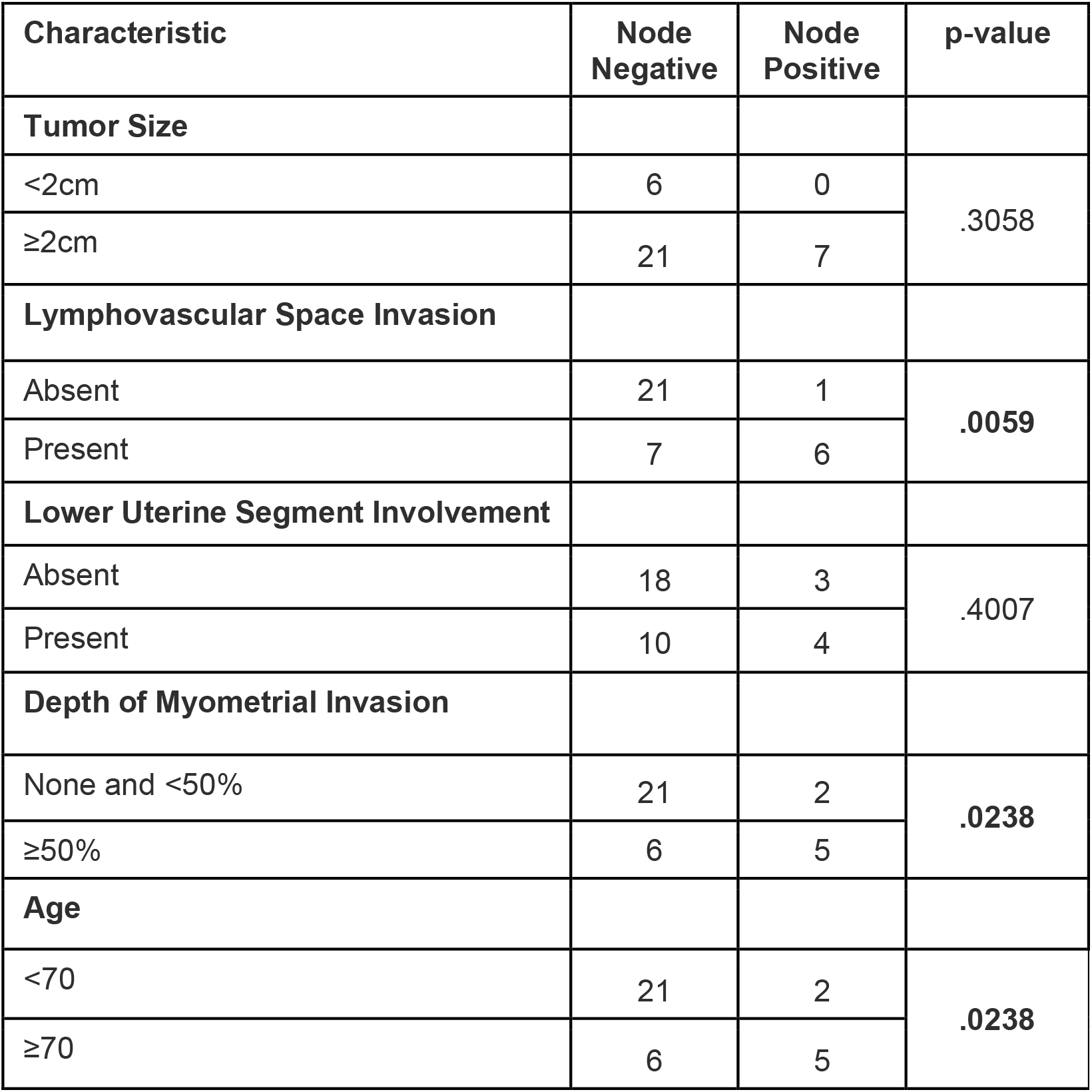
Risk factors for lymph node metastasis.

No ureteral injuries were noted. 3/35 (9%) patients experienced numbness along the distribution of the genitofemoral nerve were noted which all resolved on follow up. 5/35 asymptomatic lymphoceles were observed that did not require any further intervention. 8/35 (23%) of the patients developed symptomatic lymphedema of the lower extremities.

## Discussion

This study presents a single-surgeon, single-institution experience using a protocol of immediate intraoperative assessment of sentinel lymph nodes by frozen section [12]. Our results demonstrate 100% sensitivity and 0% false-negative rate using this protocol for high-risk endometrial histologies and add to the growing evidence supporting SLN injection and biopsy as substitute for routine complete lymphadenectomy in this patient population [7-11]. Our series confirms the accuracy of our previously published protocol for immediate intraoperative assessment of sentinel lymph node by frozen section in detecting sentinel lymph node metastases. The accuracy of this protocol is essential for high-risk endometrial cancers where the rate of lymph node metastases is elevated; here, we report a 20.6% sentinel lymph node positivity rate compared to 7.6% in our previous series which comprised predominantly low-risk endometrial cancers.

Metastatic non-sentinel lymph nodes were found in about 40% of our patients with positive sentinel lymph nodes, all of which included positive para-aortic non-sentinel lymph nodes. In the current NCCN guidelines, para-aortic lymph node dissection in endometrial cancers is left up to the discretion of the attending surgeon with little data to support this decision, particularly in high-risk histologies [14].

In vulvar cancer, a completion lymphadenectomy is recommended once macro-metastatic disease is discovered in a sentinel lymph node [15]. In breast cancer, it is recommended to perform a completion axillary lymphadenectomy when more than 2 sentinel lymph nodes are involved or no radiation planned [16, 17]. For high-risk endometrial cancers, however, similar recommendations do not exist. Randomized trials examining the utility of lymphadenectomy in endometrial cancer included mostly patients with low-risk and few with high-risk histologies [3, 4]. Several retrospective studies denote a survival benefit from completion lymphadenectomy in high-grade histologies [18-20]. Intraoperative frozen section may help determine which patients benefit from completion lymphadenectomy at the time of original surgery as well as confirm the presence of lymph node tissue in the specimen [12, 21] thus avoiding the potential morbidity of additional surgery as well as incomplete prognostic information.

The limitations of this study are that this series is a small retrospective single-surgeon and single-institutional study. Also, intraoperative assessment of sentinel lymph node protocol by frozen section may not be universally feasible or reproducible.

## Conclusion

In summary, we determined that our intraoperative SLN protocol shows 100% sensitivity and 0% false negative rate. Intraoperative SLN assessment in high-risk endometrial cancers provides immediate feedback on the success of SLN mapping and may help inform the surgeon’s decision whether to proceed with a complete lymph node dissection including pelvic and para-aortic lymph nodes.

## Data Availability

The authors confirm that the data supporting the findings of this study are available within the article [and/or] its supplementary material.

## Compliance with Ethical Standards

### Ethical approval

All procedures performed in studies involving human participants were in accordance with the ethical standards of the institutional and/or national research committee and with the 1964 Helsinki declaration and its later amendments or comparable ethical standards. Informed consent was obtained from all individual participants included in the study.

## Figures and Tables

**Table 1:** Pathological features and postoperative stage of the 35 patients who received complete study intervention, i.e. injection of dye, attempted sentinel lymph node mapping and complete surgical staging. Data are median (range) or n (%).

**Table 2:** Surgical results in patients with lymphadenectomy of the 35 patients who received complete study intervention, i.e. injection of dye, attempted sentinel lymph node mapping and complete surgical staging. Data are n (%) or median (range).

**Table 3:** Numbers and locations of positive SLN/successfully mapped SLN.

**Table 4:** 2×2 table providing sensitivity and specificity data.

**Table 5:** Risk factors for lymph node metastasis. P-values for chi-square analysis were calculated using Fisher’s exact test for 2 × 2 tables with two-tailed significance set to p < 0.05.

**Suppl. Table 1:**
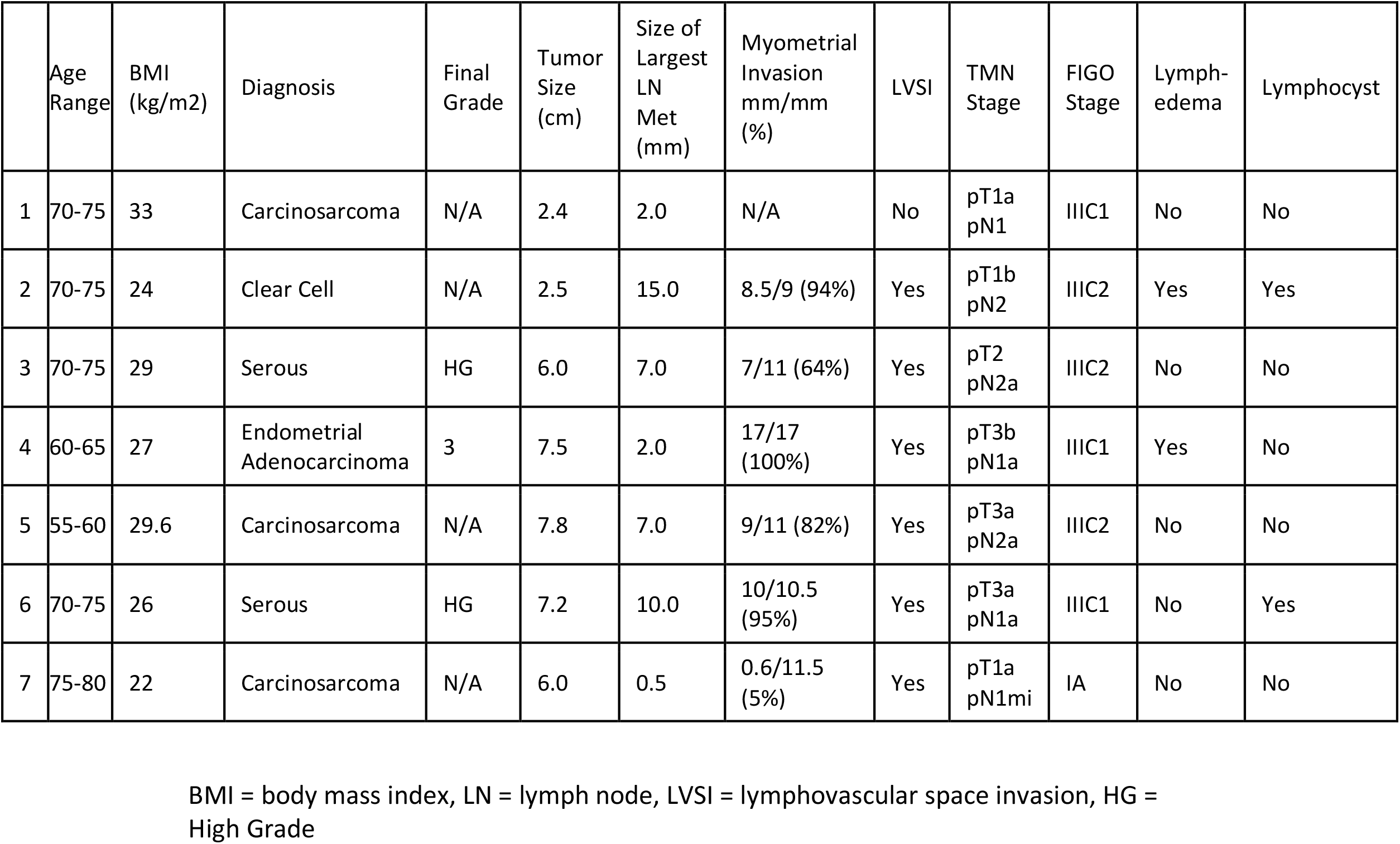
Clinical and histological characteristics of patients with positive SLN and post-operative complications

**Suppl. Table 2:** Anatomical location of positive SLNs

|  | Right |  |  | Left |  |  | Total LN | +SLN sites |
| --- | --- | --- | --- | --- | --- | --- | --- | --- |
|  | Pelvic NSLN | PA NSLN | SLN | Pelvic NSLN | PA NSLN | SLN |  |  |
| 1 | 0/12 | 0/0 | 1/5 | 0/13 | 0/0 | 0/2 | 1/32 | REI |
| 2 | 0/7 | 1/3 | 0/0 | 0/7 | 2/14 | 1/1 | 4/32 | LEI |
| 3 | 1/14 | 0/3 | 1/1 | 0/22 | 2/4 | 0/0 | 4/44 | REI |
| 4 | 0/8 | 0/3 | 2/3 | 0/6 | 0/4 | 1/3 | 3/27 | LO, RO, RO |
| 5 | 0/6 | 0/5 | 1/2 | 1/8 | 1/2 | 0/0 | 3/23 | RO |
| 6 | 0/6 | 0/2 | 1/2 | 0/10 | 0/1 | 1/2 | 4/23 | LEI, RO |
| 7 | 0/8 | 0/0 | 1/2 | 0/4 | 0/0 | 0/1 | 1/15 | REI |

NSLN = non-sentinel lymph node, PA = para-aortic, SLN = sentinel lymph node, REI = right external iliac, LEI = left external iliac, LO = left obturator, RO = right obturator

## References

1. Creasman, W.T., et al., Surgical pathologic spread patterns of endometrial cancer. A Gynecologic Oncology Group Study. Cancer, 1987. 60(8 Suppl): p. 2035–41.

2. Mikuta, J.J., International Federation of Gynecology and Obstetrics staging of endometrial cancer 1988. Cancer, 1993. 71(4 Suppl): p. 1460–3.

3. Benedetti Panici, P., et al., Systematic pelvic lymphadenectomy vs. no lymphadenectomy in early-stage endometrial carcinoma: randomized clinical trial. J Natl Cancer Inst, 2008. 100(23): p. 1707–16.

4. group, A.s., et al., Efficacy of systematic pelvic lymphadenectomy in endometrial cancer (MRC ASTEC trial): a randomised study. Lancet, 2009. 373(9658): p. 125–36.

5. Holloway, R.W., et al., Sentinel lymph node mapping and staging in endometrial cancer: A Society of Gynecologic Oncology literature review with consensus recommendations. Gynecol Oncol, 2017. 146(2): p. 405–415.

6. Renz, M., et al., Sentinel lymph node biopsies in endometrial cancer - practice patterns among Gynecologic Oncologists in the United States. J Minim Invasive Gynecol, 2019.

7. Papadia, A., et al., Retrospective validation of the laparoscopic ICG SLN mapping in patients with grade 3 endometrial cancer. J Cancer Res Clin Oncol, 2018. 144(7): p. 1385–1393.

8. Touhami, O., et al., Performance of sentinel lymph node (SLN) mapping in high-risk endometrial cancer. Gynecol Oncol, 2017. 147(3): p. 549–553.

9. Tanner, E.J., et al., The Utility of Sentinel Lymph Node Mapping in High-Grade Endometrial Cancer. Int J Gynecol Cancer, 2017. 27(7): p. 1416–1421.

10. Soliman, P.T., et al., A prospective validation study of sentinel lymph node mapping for high-risk endometrial cancer. Gynecol Oncol, 2017. 146(2): p. 234–239.

11. Ehrisman, J., et al., Performance of sentinel lymph node biopsy in high-risk endometrial cancer. Gynecol Oncol Rep, 2016. 17: p. 69–71.

12. Renz, M., et al., Immediate intraoperative sentinel lymph node analysis by frozen section is predictive of lymph node metastasis in endometrial cancer. J Robot Surg, 2019.

13. Group, G.O., Surgical Procedures Manual. www.gogmember.gog.org, revised 2005.

14. Network, N.C.C., NCCN Clinical Practice Guidelines in Oncology - Uterine Neoplasms. 2020.

15. (NCCN), N.C.C.N., NCCN Clinical Practice Guidelines in Oncology - Vulvar Cancer (Squamous Cell Carcinoma). 2020.

16. Giuliano, A.E., et al., Locoregional recurrence after sentinel lymph node dissection with or without axillary dissection in patients with sentinel lymph node metastases: the American College of Surgeons Oncology Group Z0011 randomized trial. Ann Surg, 2010. 252(3): p. 426–32; discussion 432-3.

17. Network, N.C.C., NCCN Clinical Practice Guidelines in Oncology - Breast Cancer. 2020.

18. Mahdi, H., et al., Prognostic impact of lymphadenectomy in uterine serous cancer. BJOG, 2013. 120(4): p. 384–91.

19. Mahdi, H., D. Lockhart, and M. Moselmi-Kebria, Prognostic impact of lymphadenectomy in uterine clear cell carcinoma. J Gynecol Oncol, 2015. 26(2): p. 134–40.

20. Todo, Y., et al., Survival effect of para-aortic lymphadenectomy in endometrial cancer (SEPAL study): a retrospective cohort analysis. Lancet, 2010. 375(9721): p. 1165–72.

21. Casarin, J., et al., Frozen Section for Detection of Lymph Nodes After Cervical Injection with Indocyanine Green (ICG) for Sentinel Lymph Node Technique in Endometrial Cancer Staging. Ann Surg Oncol, 2018. 25(12): p. 3692–3698.

